# The effect of yoga and aerobic exercise on children’s physical activity in rural India: a randomized controlled trial

**DOI:** 10.1101/2024.10.06.24314980

**Authors:** Tarun Reddy Katapally, Jamin Patel, Sheriff Tolulope Ibrahim, Sonal Kasture, Anuradha Khadilkar, Jasmin Bhawra

**Affiliations:** DEPtH Lab, Faculty of Health Sciences, Western University, London, Ontario, Canada; Department of Epidemiology and Biostatistics, Schulich School of Medicine and Dentistry, Western University, London, Ontario, Canada N6A 3K7; Children’s Health Research Institute, Lawson Health Research Institute, 750 Base Line Road East, Suite 300, London, Ontario, Canada N6C 2R5; Hirabai Cowasji Jehangir Medical Research Institute, Pune, Maharashtra, India; CHANGE Research Lab, School of Occupational and Public Health, Toronto Metropolitan University, Toronto, Ontario, Canada

**Author notes:** **Corresponding author:** Tarun Reddy Katapally, Ph.D., DEPtH Lab, Faculty of Health Sciences, Western University, 1151 Richmond St, London, ON N6A 5B9, London, Ontario, Canada.

**Keywords:** aerobic exercise, children’s health, global south, physical activity, yoga

## Abstract

**Purpose:** The objective of this study was to test the effect of both yoga and aerobic exercise on children’s moderate-to-vigorous physical activity (MVPA) in rural India.

**Methods:** The study utilized secondary data from a randomized, controlled, open-labelled, single-center, two-site, parallel-group trial. The study was conducted in rural India over a 6-month period between 2018-2019. Children aged 6 to 11 years were randomized into three groups: aerobic exercise (30 minutes, 5 days/week), control (no intervention), and yoga (30 minutes, 5 days/week). MVPA was measured at baseline and at six months using the Quantification of Physical Activity in School Children and Adolescents survey adapted and validated for Indian children. Overall sample and gender-segregated data were analyzed using paired sample t-tests and one-way analysis of variance with post-hoc analyses.

**Findings:** In the overall sample (N=151), mean MVPA (minutes/day) increased significantly in both yoga (n=50; p<0.001) and aerobic exercise (n=49; p<0.001) groups from baseline to endline. Among males, mean MVPA increased significantly from baseline to endline in all three groups, including the control group (n=23; p=0.005). Among females, mean MVPA increased only in the yoga group, with baseline to endline change being significant across the three groups (p=0.005), and with the yoga group depicting greater change in comparison to the control group (p=0.004).

**Conclusions:** Our findings suggest that both yoga and aerobic exercise can increase MVPA among rural children, with yoga being particularly beneficial for girls – a significant finding to inform culturally-appropriate active living policies to minimize the current physical activity gender gap in India. These findings can have implications for public health programs and policies not only in India but across other rural areas worldwide, where similar challenges in promoting physical activity among children may exist.

## 1. Introduction

Physical inactivity is a major contributor to global non-communicable diseases (NCD), [1–3] with an estimated mortality burden of 3.2 million deaths per year [4]. Physical inactivity is a significant concern in pediatric populations as it has been linked to the prevalence of NCDs in adulthood [5– 7]. Between 2020 and 2030, the cumulative economic cost of India’s physical inactivity is projected to be USD 35.4 billion [6]. Evidence indicates that physical activity among children and youth in India has consistently decreased since 2016, especially during the coronavirus disease pandemic.[8–10] This decline in physical activity poses a critical challenge for pediatric healthcare providers and policymakers in promoting and facilitating active lifestyles among children [11–13].

Child and youth physical activity is below recommended guidelines [8–10] despite growing policies and programs [10], which range from school educational curricula and governmental investments in sports infrastructure [10] to comprehensive sports programs [14], and physical education interventions across India [15,16]. Culturally-appropriate strategies, which consider the identity and heritage of global south populations need to be considered seriously to reverse this growing physical inactivity trend [10,17–19]. Given its historical and cultural significance, yoga has the potential to serve as a complementary medicine practice to promote physical activity among Indian children in particular [10,19,20]. Both breathing (pranayama), and physical techniques (asanas) [21] have been shown to improve motor function [22–24], muscle strength [22,25], and mental health [22,26,27] among Indian children and youth.

While the benefits of yoga are well-established, irrespective of the type of yoga practice [28], currently there is no concrete evidence linking yoga with moderate to vigorous physical activity (MVPA) among children and youth – a key indicator informing global physical activity guidelines [29,30]. Some preliminary correlational evidence is being explored [20], however; it is important to establish empirical evidence via randomized controlled trials to understand these relationships better. We hypothesize that while the practice of yoga itself can result in the accumulation of more MVPA [31], yoga practice can inherently influence other active living behaviours [32–34], which might enhance MVPA. However, it is critical to test this hypothesis in parallel with the effect of aerobic exercise on MVPA, with the obvious hypothesis being that aerobic exercise would increase MVPA [5,29]. It is also important to understand gender variations of the impact of yoga on MVPA to inform healthcare strategies, policies, and programs that promote equitable promotion of MVPA among girls in India – who are generally less physically active than Indian boys [9,10]. Thus, as part of a randomized controlled study, our objective is to investigate the influence of yoga and aerobic exercise on MVPA of children in rural India, while exploring gender variations of this effect.

## 2. Methods

### 2.1 Study design and setting

This study is a secondary analysis of a randomized, controlled, open-labelled, single-centre, 2-site, three-arm (allocation ratio: 1:1:1) parallel-group trial of yoga, aerobic exercise, and protein supplementation conducted among 232 children (aged 6 to 11 years) [25]. Children were recruited in two randomly selected government schools in two villages located 60km from Pune, India from July 13^th^ to 25^th^ 2018. Parents provided written informed consent, while their children provided written assent. Ethics approval was obtained from the Hirabai Cowasji Jehangir Medical Research Institute Ethics Review Committee (ECR/352/Inst/MH/2013/RR-16). The study was registered with the Clinical Trials Registry - India (CTRI/2018/07/014815). **Figure 1** displays the recruitment, randomization, and group allocation process to determine the final sample size. Additional methodological information, including sample size calculations, has been published by Kasture et al (2023) [25].

**Figure 1.**
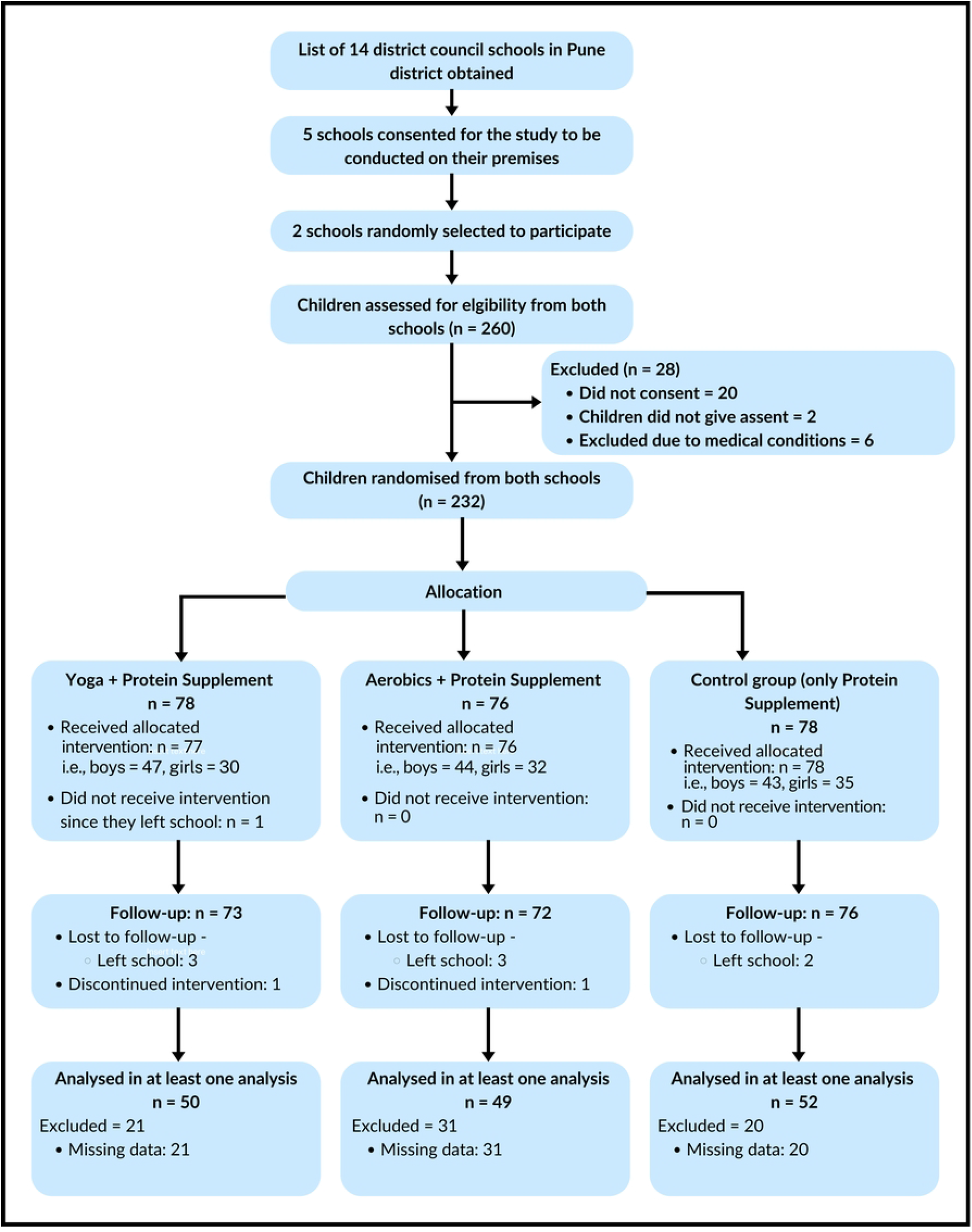
CONSORT diagram depicting study design and participant flow.

### 2.2 Inclusion and Exclusion Criteria

Male children between the ages of 7 and 11 years and female children between the ages of 6 and 10 years were included as evidence indicates that female children attain puberty sooner [35,36]. Additionally, children had to provide MVPA data at baseline and endline to be included in the analyses of this study. Children using medication known to affect bone/muscle health or with chronic illnesses were excluded from the study [25].

### 2.3 Recruitment and Randomization

Before enrolment, parents of children (N = 260) who met the age criteria were approached. Only 240 parents and their children provided written informed consent and assent, respectively. To achieve randomization, a data manager who was not involved in data collection generated a random sequence via a web-based application [37] to randomize children into three mutually exclusive groups (yoga, aerobic exercise, and control).

### 2.4 The Six-Month Intervention

All children across the three randomized groups (control, yoga, and aerobic exercise) received a protein supplement (Ladoo) for 6 days a week over six months. The ingredients for the protein supplement were roasted groundnuts, roasted sesame seeds, fresh dates, skimmed milk powder and powdered cane sugar [25]. Children in the yoga group engaged in a 30-minute, five-day-a-week yoga exercise (**Figure 2**) before school. This exercise was a type of yoga called Yogasanas, which involved engagement in various static poses. The poses required children to use their major joint and muscle groups, including their trunk, back, hip and knee muscles.

**Figure 2.**
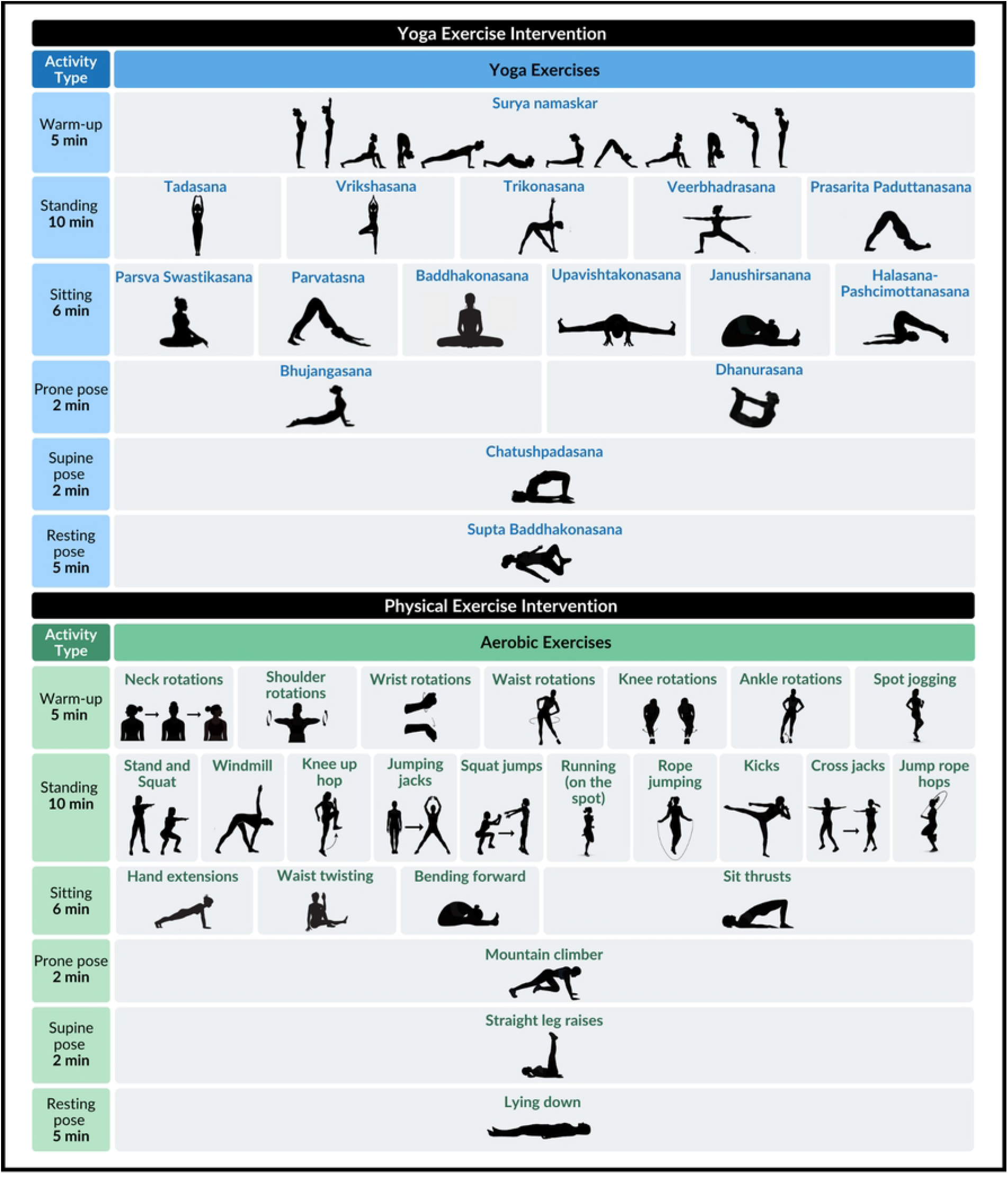
Daily yoga and aerobic exercise intervention regimen

Children in the aerobic group engaged in aerobic exercise for 30 minutes, five days a week (**Figure 2**). Daily physical exercise involved a five-minute warm-up, a 20-minute core and lower limb strengthening exercise, and a five-minute cool-down period. In week one, instructors provided hands-on support and visual demonstration to ensure correct posture for children in both groups. Thereafter, visual demonstration and verbal guidance were used to engage with children.

### 2.5 Data Collection

Data collection was obtained both at baseline (before randomization) and endline (after 6 months). Age, gender, body mass index (BMI), sunlight exposure, dietary intake data, as well as data on the primary outcome variable, MVPA, were obtained via researcher-administered validated surveys in the presence of children’s caregivers. The Quantification of Physical Activity in School Children and Adolescents survey was adapted and validated for Indian children before being used to obtain MVPA data [38,39]. Anthropometry and body composition, including height and weight, were measured using bioelectrical impedance analysis Tanita Body Composition Analyzer [Model BC-420MA].

### 2.6 Data analysis

All statistical analyses were conducted in R 4.2.1 [40]. Mean and standard deviation were used to describe age, BMI, gender, and minutes of MVPA/day. Paired sample t-tests assessed differences in mean MVPA/day within the yoga, aerobic, and control groups from pre-to post-intervention. Mean differences (MD), defined as the difference in mean MVPA/day between pre-and post-intervention, were calculated for the three groups. Analysis of variance with post hoc Tukey tests was used to detect differences in baseline characteristics and MVPA at both pre-and post-intervention, as well as MD between the three groups. Data analyses were segregated by gender (male vs. female). Results were deemed statistically significant at p<0.05.

## 3. Results

A total of 151 (N) children were included in the final analyses, with the following distribution across the three groups: yoga (n=50), aerobic exercise (n=49), and control (n=52). The sample comprised of 54.30% males and 45.70% females. **S1 Table** depicts no statistically significant differences in the distribution of age and gender between the three groups, both at baseline and endline. However, there was a statistically significant difference in BMI across the three groups at endline (p=0.015). The mean BMI was 14 at baseline and 14.38 at endline, with children in the aerobic exercise group having higher body mass index at both time points.

**S2 Table** depicts pre- (baseline) and post-intervention (endline) mean MVPA/day differences within and across the three groups (yoga vs aerobic vs control). These differences are shown in the overall sample, as well as in the gender-segregated samples (males and females). The mean MVPA/day in the overall sample was 54.02 minutes/day and 80.15 minutes/day at baseline and endline, respectively. In the overall sample, mean MVPA/day increased significantly between baseline and endline within both yoga [mean difference (MD) = 35.85, 95% confidence interval (CI) = 18.77, 52.92] and aerobic exercise groups [MD=32.21, 95% CI=18.15,46.27]. There was no significant difference in mean MVPA/day between yoga, aerobic and control groups either at baseline or endline; however, the yoga group depicted higher MD from baseline to endline (p=0.057).

Among males, the mean MVPA/day was 57.53 minutes/day and 98.01 minutes/day at baseline and endline, respectively. Moreover, mean MVPA/day among males increased from baseline to endline among all three groups: yoga [MD=35.51, 95% CI=11.47, 59.56], aerobic exercise [MD=45.97, 95% CI=31.64, 60.31], and control [MD=40.51, 95% CI=13.05, 67.96]. There was no significant difference in mean MVPA/day between the three groups at baseline or endline; however, the aerobic group showed higher MD from baseline to endline (p=0.779). Among females, the mean MVPA/day was 49.86 minutes/day and 58.93 minutes/day at baseline and endline, respectively. More importantly, among females, mean MVPA/day increased significantly between baseline and endline only within the yoga group [MD=36.40, 95% CI=11.52, 61.26], with MD of MVPA from baseline to endline being significant not only across the three groups (p=0.005) but also being higher among the yoga group in comparison to control (p=0.004) and aerobic exercise (p=0.328) groups. **Figure 3** shows three sets of boxplots and density plots comparing the distribution of MVPA/day at both baseline and endline across all three groups segregated by overall, male, and female samples. In the overall sample, across all three groups, the distribution of MVPA/day was skewed at baseline, but the skewness was reduced at endline. In both gender-segregated samples, a similar skewness in the distribution of MVPA/day was observed in both yoga and aerobic exercise groups at baseline. Nevertheless, post-intervention (i.e., at endline), the skewness in the distribution of MVPA/day reduced – similar to the overall sample, in both males and females.

**Figure 3.**
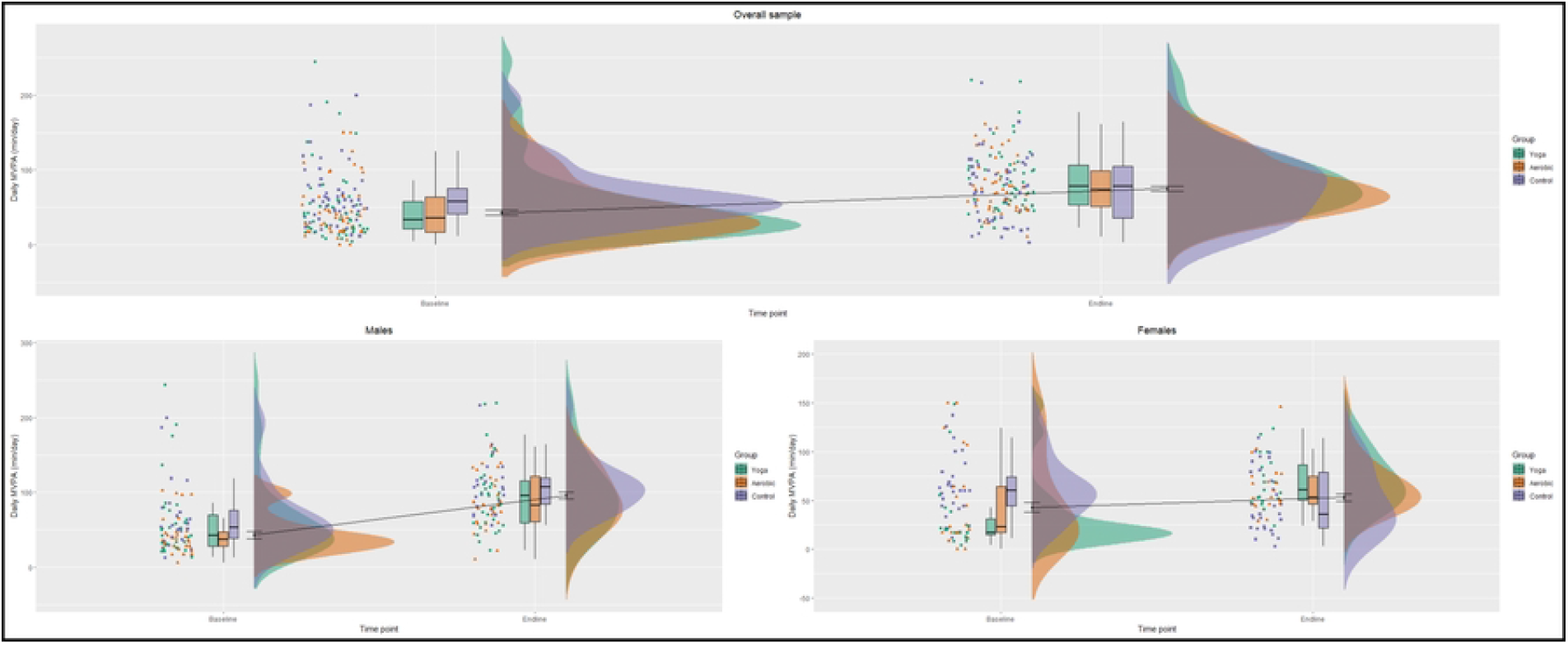
Changes in MVPA/day distribution from baseline to endline. Yoga: green, aerobic exercise: orange, control: purple.

## 4. Discussion

This study explored the effect of both yoga and aerobic exercise on children’s MVPA. The findings showed that both yoga and aerobic exercise, when consistently incorporated into a randomized controlled trial over a six-month period, led to statistically significant increases in MVPA levels among rural children aged 6-11 years. It is also important to note that children who did not receive either the yoga or aerobic exercise intervention (control group) did not show a statistically significant increase in their MVPA levels between baseline and endline. However, there were no differences in pre/post-intervention MVPA levels between the yoga and aerobic exercise groups. Nevertheless, more nuanced findings were observed after conducting gender-segregated analyses. Among males, MVPA increased significantly from baseline to endline in all three groups, including the control group. Among females, MVPA increased from baseline to endline only in the yoga group, with the change in mean MVPA from baseline to endline being significantly higher in the yoga group in comparison to the control group.

The benefits of aerobic exercise interventions on MVPA levels among children are well established [5,41,42], and this study adds novel insights to the existing evidence by examining the effect of aerobic exercise on rural Indian children. While there is an emerging body of evidence – particularly the use of randomized control trials – to assess the effects of yoga on physical fitness and mental health [22,26,43–47], a gap in evidence remains regarding the effect of yoga practice on the overall accumulation of MVPA among children. Thus far, to our knowledge, no study has examined the impact of yoga practice on the MVPA levels of children, who are at a critical stage both in terms of physiological development as well as the formation of active living behaviours.

There is some evidence from cross-sectional studies showing that yoga is associated with higher levels of MVPA in adult populations [34]. Among children, studies have primarily focused on the general health benefits of yoga, such as improvements in mental health [22,26,27], flexibility [48], and stress reduction [27,49]. This study, however, provides novel insights by linking yoga practice directly to increased MVPA levels, a key indicator of health [29] and the primary measure that informs global guidelines on the physical of children and youth [30]. More importantly, our findings reinforce the findings of a cross-sectional study conducted among children and youth across 28 cities and villages in India, which found that yoga practice was associated with more minutes of MVPA per day [20].

Another critical finding is that yoga can be a culturally-appropriate alternative or supplement to aerobic exercise, particularly among Indian children and youth [10,20,25]. The potential to implement yoga as part of physical activity programming is a significant finding because physical inactivity among children and youth in India has consistently increased since 2016 [9,10], despite growing active living policies and programs [10]. It is time to stem this growing physical inactivity trend by incorporating culturally appropriate and accessible practices such as yoga into routine pediatric care, health promotion strategies, and school health interventions [10,17,18,25].

Yoga, as an alternative to aerobic exercise, might be particularly relevant to rural areas, where resources and physical activity infrastructure in schools may be limited [50]. As yoga is not only culturally relevant but also cost-effective and easy to implement [25], there is potential to incorporate its practice into public health strategies [51]. Moreover, promoting yoga leverages existing cultural knowledge and practices [10], potentially enhancing community engagement [10,51] and support for physical activity programs [20,52]. As an ancient Indian practice, the incorporation of culturally-appropriate exercises is also relevant in the context of decolonizing physical activity [53–57], which requires a holistic approach that integrates health and education systems [58]. Decolonizing active living entails providing children with diverse physical activity options which recognize their local heritage and culture – which vary widely across the Indian subcontinent i.e., can go beyond just yoga to include dance, and martial arts, among other practices [53,55,59]. Nonetheless, as our findings do not indicate a difference between aerobic exercise and yoga in improving overall MVPA among children, it is important to note that we do not propose yoga as a replacement for aerobic exercise – which has immense benefits to children’s health [5,22,42] – rather it is a practice which warrants consideration for physical activity programs in the Indian context.

The findings also indicate gender-specific effects, with MVPA levels increasing from baseline to endline in both the yoga and aerobic exercise groups among males, while MVPA levels increasing only in the yoga group among females. Perhaps most importantly, this change in MVPA from baseline to endline was significant but not in the control group - suggesting that yoga may be particularly effective for increasing physical activity levels among female children. This finding has particular implications in mitigating and/or minimizing barriers to physical activity accumulation commonly experienced among girls due to cultural norms and societal expectations, particularly in rural India [9,10]. These findings underscore the importance of considering gender when designing active living policies and healthcare strategies [60], as gender-specific strategies are necessary to ensure active living equity [10,61,62].

Interestingly, when the distribution of MVPA was explored across the three samples (overall, males, and females), we found that the baseline distribution of MVPA levels was skewed across all three groups, with most children engaging in low levels of MVPA. However, in the yoga and aerobic groups, the MVPA levels not only increased, but the distribution became more normally distributed at the six-month follow-up. This indicates that children who were not engaging in sufficient physical activity at baseline not only showed an increase in their MVPA at endline, but also that the MVPA levels were more evenly distributed (i.e., similar). This observation has implications for prompting equity in physical activity to ensure that all children, regardless of their starting point, have the opportunity to improve their health outcomes [51,63,64].

This study found that both yoga and aerobic exercise interventions increased MVPA levels among rural Indian children. Apart from yoga practice and aerobic exercise causing exertion that increases MVPA accumulation, yoga or aerobic exercise also promotes overall physical fitness [22,25] and health [7,29,51,65], which naturally leads to an increase in physical activity levels [22,66]. Moreover, yoga has been shown to improve muscle function [22,25], motor function [22–24], and cardiorespiratory fitness [22] which can enable children to participate in a broad range of physical activities with greater ease [67].

While both yoga and aerobic exercise might contribute to higher MVPA levels, yoga might be especially pertinent in India not only because it is culturally relevant, but also because it can be practiced in low-resource settings [25,52,68,69]. This makes yoga accessible to a wider population, particularly in rural regions where active living infrastructure is limited [50,63,70,71]. Additionally, yoga can be practiced indoors in all weather conditions, an often overlooked component in promoting active living to counter air pollution, regardless of cultural, built environment, gender, and safety barriers [8–10] which influence MVPA levels among children and youth [72–74].

## 5. Strengths and limitations

The randomized controlled design enables a robust comparison between intervention groups and by minimizing potential biases. The inclusion of a culturally appropriate intervention such as yoga, provides novelty and relevance, particularly in developing relevant active living policies for children and youth in India. Moreover, conducting the study in rural Indian schools helps address another gap in evidence [8–10]. Furthermore, the gender-stratified analyses add another layer of strength, providing insights into how tailored active living strategies and practices can be developed to address the gender gap in active living [8–10]. A limitation of this study is the reliance on self-reported MVPA data, which can be subject to recall bias and inaccuracies, potentially affecting the reliability of the measurements.

Additionally, the study was conducted in only two district schools, which may limit the generalizability of the findings to other regions or populations. Finally, while the six-month follow-up period provides valuable insights into the medium-term effects, it does not capture the long-term sustainability of the interventions. Future studies should be conducted longitudinally over a longer period and with larger samples across the diverse geographic regions in India to capture the impact of yoga and aerobic exercise on the MVPA of children.

## 6. Conclusions

To our knowledge, this is the first study to examine the impact of yoga and aerobic exercise on MVPA levels of children. This study demonstrates that both yoga and aerobic exercise can significantly increase MVPA levels among rural Indian children, highlighting their effectiveness in promoting physical activity in resource-limited settings. The findings emphasize the value of culturally-appropriate interventions like yoga, particularly to minimize the gender gap, and enable equity in active living – findings that can have implications for active living policies and practices not only in India but across other global south countries that can potentially benefit from decolonizing active living strategies.

## Data Availability

The raw data required to reproduce the above findings are available to download from: https://doi.org/10.6084/m9.figshare.25970053.v1

## S1 Appendix

Change in average MVPA of children from baseline to endline.

Yoga intervention group: green; aerobic intervention group: orange; and control group: purple. First panel: overall sample; second panel: male subsample; and third panel: female subsample.

## Acknowledgements

We acknowledge Hirabai Cowasji Jehangir Medical Research Institute for their support in obtaining this valuable data, and Heya Desai for her contributions to visualization and collation of this manuscript. Ethics approval was obtained from Hirabai Cowasji Jehangir Medical Research Institute Ethics Review Committee (ECR/352/Inst/MH/2013/RR-16).

## References

1. Katzmarzyk PT, Friedenreich C, Shiroma EJ, Lee IM. Physical inactivity and non-communicable disease burden in low-income, middle-income and high-income countries. Br J Sports Med. 2022 Jan 1;56(2):101–6.

2. Lee IM, Shiroma EJ, Lobelo F, Puska P, Blair SN, Katzmarzyk PT. Effect of physical inactivity on major non-communicable diseases worldwide: an analysis of burden of disease and life expectancy. The Lancet. 2012 Jul 21;380(9838):219–29.

3. Santos AC, Willumsen J, Meheus F, Ilbawi A, Bull FC. The cost of inaction on physical inactivity to public health-care systems: a population-attributable fraction analysis. Lancet Glob Health. 2023 Jan 1;11(1):e32–9.

4. Shimizu Y. Physical activity [Internet]. Health topics. [cited 2024 May 6]. Available from: https://www.who.int/westernpacific/health-topics/physical-activity

5. Bull FC, Al-Ansari SS, Biddle S, Borodulin K, Buman MP, Cardon G, et al. World Health Organization 2020 guidelines on physical activity and sedentary behaviour. Br J Sports Med. 2020 Dec 1;54(24):1451–62.

6. World Health Organization. Physical activity India 2022 country profile [Internet]. Physical activity India 2022 country profile. 2022 [cited 2024 May 12]. Available from: https://www.who.int/publications/m/item/physical-activity-ind-2022-country-profile

7. Poitras VJ, Gray CE, Borghese MM, Carson V, Chaput JP, Janssen I, et al. Systematic review of the relationships between objectively measured physical activity and health indicators in school-aged children and youth. Appl Physiol Nutr Metab. 2016 Jun;41(6 (Suppl. 3)):S197–239.

8. Katapally TR, Goenka S, Bhawra J, Mani S, Krishnaveni GV, Kehoe SH, et al. Results From India’s 2016 Report Card on Physical Activity for Children and Youth. J Phys Act Health. 2016 Nov 1;13(2):S176–82.

9. Bhawra J, Chopra P, Harish R, Mohan A, Ghattu KV, Kalyanaraman K, et al. Results from India’s 2018 Report Card on Physical Activity for Children and Youth. J Phys Act Health. 2018 Jan 2;15(2):S373–4.

10. Bhawra J, Khadilkar A, Krishnaveni GV, Kumaran K, Katapally TR. The 2022 India Report Card on physical activity for children and adolescents. J Exerc Sci Fit. 2023 Jan;21(1):74– 82.

11. Lobelo F, Muth ND, Hanson S, Nemeth BA, COUNCIL ON SPORTS MEDICINE and FITNESS, SECTION ON OBESITY, et al. Physical Activity Assessment and Counseling in Pediatric Clinical Settings. Pediatrics. 2020 Mar 1;145(3):e20193992.

12. Pellerine LP, O’Brien MW, Shields CA, Crowell SJ, Strang R, Fowles JR. Health Care Providers’ Perspectives on Promoting Physical Activity and Exercise in Health Care. Int J Environ Res Public Health. 2022 Jan;19(15):9466.

13. Muth ND, Bolling C, Hannon T, Sharifi M, SECTION ON OBESITY, COMMITTEE ON NUTRITION. The Role of the Pediatrician in the Promotion of Healthy, Active Living. Pediatrics. 2024 Feb 26;153(3):e2023065480.

14. Khelo India [Internet]. Khelo India. 2022 [cited 2024 May 13]. Available from: https://kheloindia.gov.in/index.html

15. Sportz Village Schools - In-School Sports Program [Internet]. Sportz Village Schools – India’s No.1 Sports Education Organization. [cited 2024 May 13]. Available from: https://edusports.sportzvillage.com/in-school-sports-program/

16. Fit India - Be fit [Internet]. [cited 2024 Jun 1]. Available from: https://fitindia.gov.in/

17. Joo JY, Liu MF. Culturally tailored interventions for ethnic minorities: A scoping review. Nurs Open. 2021;8(5):2078–90.

18. Horne M, Tierney S, Henderson S, Wearden A, Skelton DA. A systematic review of interventions to increase physical activity among South Asian adults. Public Health. 2018 Sep 1;162:71–81.

19. Leach MJ, Veziari Y, Flanagan C, Schloss J. Prevalence of Complementary Medicine Use in Children and Adolescents: A Systematic Review. J Pediatr Health Care [Internet]. 2024 Jan 22 [cited 2024 Jun 29];0(0). Available from: https://www.jpedhc.org/article/S0891-5245(23)00372-3/fulltext

20. Patel J, Ibrahim S, Bhawra J, Khadilkar A, Katapally TR. Association between yoga and related contextual factors with moderate-to-vigorous physical activity among children and youth aged 5 to 17 years across five Indian states. PeerJ. 2024 May 31;12:e17369.

21. Govindaraj R, Karmani S, Varambally S, Gangadhar BN. Yoga and physical exercise – a review and comparison. Int Rev Psychiatry. 2016 May 3;28(3):242–53.

22. Telles S, Singh N, Bhardwaj AK, Kumar A, Balkrishna A. Effect of yoga or physical exercise on physical, cognitive and emotional measures in children: a randomized controlled trial. Child Adolesc Psychiatry Ment Health. 2013 Nov 7;7(1):37.

23. Folleto JC, Pereira KR, Valentini NC. The effects of yoga practice in school physical education on children’s motor abilities and social behavior. Int J Yoga. 2016;9(2):156–62.

24. Barnett LM, Verswijveren Sjjm, Colvin B, Lubans DR, Telford RM, Lander NJ, et al. Motor skill competence and moderate- and vigorous-intensity physical activity: a linear and non-linear cross-sectional analysis of eight pooled trials. Int J Behav Nutr Phys Act. 2024 Feb 7;21(1):14.

25. Kasture S, Khadilkar A, Padidela R, Gondhalekar K, Patil R, Khadilkar V. Effect of Yoga or Physical Exercise on Muscle Function in Rural Indian Children: A Randomized Controlled Trial. J Phys Act Health. 2023 Nov 6;21(1):85–93.

26. Hart N, Fawkner S, Niven A, Booth JN. Scoping Review of Yoga in Schools: Mental Health and Cognitive Outcomes in Both Neurotypical and Neurodiverse Youth Populations. Children. 2022 Jun;9(6):849.

27. Kauts A, Sharma N. Effect of yoga on academic performance in relation to stress. Int J Yoga. 2009;2(1):39–43.

28. Cramer H, Lauche R, Langhorst J, Dobos G. Is one yoga style better than another? A systematic review of associations of yoga style and conclusions in randomized yoga trials. Complement Ther Med. 2016 Apr 1;25:178–87.

29. US Department of Health and Human Services. Youth Physical Activity Guidelines | Physical Activity | Healthy Schools | CDC [Internet]. 2022 [cited 2024 May 13]. Available from: https://www.cdc.gov/healthyschools/physicalactivity/guidelines.htm

30. WHO guidelines on physical activity and sedentary behaviour [Internet]. 2020 [cited 2024 Jun 1]. Available from: https://www.who.int/publications-detail-redirect/9789240015128

31. Larson-Meyer DE. A Systematic Review of the Energy Cost and Metabolic Intensity of Yoga. Med Sci Sports Exerc. 2016 Aug;48(8):1558.

32. Wang WL, Chen KH, Pan YC, Yang SN, Chan YY. The effect of yoga on sleep quality and insomnia in women with sleep problems: a systematic review and meta-analysis. BMC Psychiatry. 2020 May 1;20(1):195.

33. Polsgrove MJ, Eggleston BM, Lockyer RJ. Impact of 10-weeks of yoga practice on flexibility and balance of college athletes. Int J Yoga. 2016;9(1):27–34.

34. Watts AW, Rydell SA, Eisenberg ME, Laska MN, Neumark-Sztainer D. Yoga’s potential for promoting healthy eating and physical activity behaviors among young adults: a mixed-methods study. Int J Behav Nutr Phys Act. 2018 May 2;15(1):42.

35. McDowell MA, Brody DJ, Hughes JP. Has age at menarche changed? Results from the National Health and Nutrition Examination Survey (NHANES) 1999-2004. J Adolesc Health Off Publ Soc Adolesc Med. 2007 Jan 23;40(3):227–31.

36. Puberty: Stages for Boys & Girls [Internet]. Cleveland Clinic. [cited 2024 Jun 2]. Available from: https://my.clevelandclinic.org/health/articles/22192-puberty

37. RANDOM.ORG - True Random Number Service [Internet]. [cited 2024 Jun 1]. Available from: https://www.random.org/

38. Barbosa N, Sanchez CE, Vera JA, Perez W, Thalabard JC, Rieu M. A Physical Activity Questionnaire: Reproducibility and Validity. J Sports Sci Med. 2007 Dec 1;6(4):505–18.

39. Khadilkar AV, Chiplonkar SA, Kajale NA, Ekbote VH, Parathasarathi L, Padidela R, et al. Impact of dietary nutrient intake and physical activity on body composition and growth in Indian children. Pediatr Res. 2018 Apr;83(4):843–50.

40. R Core Team. R: A language and environment for statistical computing. [Internet]. R Foundation for Statistical Computing. 2021 [cited 2024 Jun 1]. Available from: https://www.R-project.org/

41. Gordon ES, Tucker P, Burke SM, Carron AV. Effectiveness of physical activity interventions for preschoolers: a meta-analysis. Res Q Exerc Sport. 2013 Sep;84(3):287–94.

42. van der Fels Imj, Hartman E, Bosker RJ, de Greeff JW, de Bruijn AGM, Meijer A, et al. Effects of aerobic exercise and cognitively engaging exercise on cardiorespiratory fitness and motor skills in primary school children: A cluster randomized controlled trial. J Sports Sci. 2020 Sep 1;38(17):1975–83.

43. Varambally S, Holla B, Venkatasubramanian G, Mullapudi T, Raj P, Shivakumar V, et al. Clinical effects of a yoga-based intervention for patients with schizophrenia — A six-month randomized controlled trial. Schizophr Res. 2024 Jul 1;269:144–51.

44. Kongkaew C, Lertsinthai P, Jampachaisri K, Mongkhon P, Meesomperm P, Kornkaew K, et al. The Effects of Thai Yoga on Physical Fitness: A Meta-Analysis of Randomized Control Trials. J Altern Complement Med. 2018 Jun;24(6):541–51.

45. Lang AJ, Malaktaris A, Maluf KS, Kangas J, Sindel S, Herbert M, et al. A randomized controlled trial of yoga vs nonaerobic exercise for veterans with PTSD: Understanding efficacy, mechanisms of change, and mode of delivery. Contemp Clin Trials Commun. 2021 Mar 1;21:100719.

46. Bieber M, Görgülü E, Schmidt D, Zabel K, Etyemez S, Friedrichs B, et al. Effects of body-oriented yoga: a RCT study for patients with major depressive disorder. Eur Arch Psychiatry Clin Neurosci. 2021;271(7):1217–29.

47. Yadav SS, Saoji AA, Somanadhapai S, Yadav N lal, Upadhyay J, Rishi NN, et al. Effect of Yoga-based breathing practices on depression, anxiety, stress, and fear of COVID-19 positive hospitalized patients: A randomized controlled trial. J Ayurveda Integr Med. 2024 Mar 1;15(2):100897.

48. The effects of yoga practice on balance, strength, coordination and flexibility in healthy children aged 10-12 years - PubMed [Internet]. 2019 [cited 2024 Jun 1]. Available from: https://pubmed.ncbi.nlm.nih.gov/31733751/

49. James-Palmer A, Anderson EZ, Zucker L, Kofman Y, Daneault JF. Yoga as an Intervention for the Reduction of Symptoms of Anxiety and Depression in Children and Adolescents: A Systematic Review. Front Pediatr [Internet]. 2020 Mar 13 [cited 2024 Jun 1];8. Available from: https://www.frontiersin.org/articles/10.3389/fped.2020.00078

50. Pfledderer CD, Burns RD, Byun W, Carson RL, Welk GJ, Brusseau TA. School-based physical activity interventions in rural and urban/suburban communities: A systematic review and meta-analysis. Obes Rev. 2021;22(9):e13265.

51. Global action plan on physical activity 2018–2030: more active people for a healthier world [Internet]. 2018 [cited 2024 Jun 1]. Available from: https://www.who.int/publications-detail-redirect/9789241514187

52. Mohanty S, Epari V, Yasobant S. Can Yoga Meet the Requirement of the Physical Activity Guideline of India? A Descriptive Review. Int J Yoga. 2020;13(1):3–8.

53. Knuth AG, Leite GS, Dos Santos SF da S, Crochemore-Silva I. Is It Possible to Decolonize the Field of Physical Activity and Health? J Phys Act Health. 2024 Apr 24;1–3.

54. Pang B, Balram R, Knijnik J. Decolonization in Sport: Reimagining Embodiment from an Anthropocosmic Perspective. In: Ravulo J, Olcoń K, Dune T, Workman A, Liamputtong P, editors. Handbook of Critical Whiteness: Deconstructing Dominant Discourses Across Disciplines [Internet]. Singapore: Sprimger Nature; 2023 [cited 2024 Jun 3]. p. 1–12. Available from: 10.1007/978-981-19-1612-0_42-1

55. Ganneri NR. The Debate on ‘Revival’ and the Physical Culture Movement in Western India (1900–1950). In: Sport Across Asia. Routledge; 2012.

56. Alter J. Subaltern Bodies and Nationalist Physiques: Gama the Great and the Heroics of Indian Wrestling. Body Soc - BODY SOC. 2000 Jun 1;6:45–72.

57. Chakraborty C. The Hindu ascetic as fitness instructor: Reviving faith in yoga. Int J Hist Sport. 2007 Sep 1;24:1172–86.

58. Katapally TR. JMIR Pediatrics and Parenting - Smart Indigenous Youth: The Smart Platform Policy Solution for Systems Integration to Address Indigenous Youth Mental Health [Internet]. 2020 [cited 2024 Jun 2]. Available from: https://pediatrics.jmir.org/2020/2/e21155/

59. Aubert S, Barnes JD, Demchenko I, Hawthorne M, Abdeta C, Nader PA, et al. Global Matrix 4.0 Physical Activity Report Card Grades for Children and Adolescents: Results and Analyses From 57 Countries. J Phys Act Health. 2022 Oct 22;19(11):700–28.

60. Camacho-Miñano MJ, LaVoi NM, Barr-Anderson DJ. Interventions to promote physical activity among young and adolescent girls: a systematic review. Health Educ Res. 2011 Dec 1;26(6):1025–49.

61. Bourke M, Haddara A, Loh A, Carson V, Breau B, Tucker P. Adherence to the World Health Organization’s physical activity recommendation in preschool-aged children: a systematic review and meta-analysis of accelerometer studies. Int J Behav Nutr Phys Act. 2023 Apr 26;20(1):52.

62. Kretschmer L, Salali GD, Andersen LB, Hallal PC, Northstone K, Sardinha LB, et al. Gender differences in the distribution of children’s physical activity: evidence from nine countries. Int J Behav Nutr Phys Act. 2023 Sep 4;20(1):103.

63. Owen KB, Nau T, Reece LJ, Bellew W, Rose C, Bauman A, et al. Fair play? Participation equity in organised sport and physical activity among children and adolescents in high income countries: a systematic review and meta-analysis. Int J Behav Nutr Phys Act. 2022 Mar 18;19(1):27.

64. Gautam N, Dessie G, Rahman MM, Khanam R. Socioeconomic status and health behavior in children and adolescents: a systematic literature review. Front Public Health [Internet]. 2023 Oct 17 [cited 2024 Jun 1];11. Available from: https://www.frontiersin.org/journals/public-health/articles/10.3389/fpubh.2023.1228632/full

65. Janssen I, LeBlanc AG. Systematic review of the health benefits of physical activity and fitness in school-aged children and youth. Int J Behav Nutr Phys Act. 2010 May 11;7(1):40.

66. Collins K, Staples K. The role of physical activity in improving physical fitness in children with intellectual and developmental disabilities. Res Dev Disabil. 2017 Oct 1;69:49–60.

67. Gäbler M, Prieske O, Hortobágyi T, Granacher U. The Effects of Concurrent Strength and Endurance Training on Physical Fitness and Athletic Performance in Youth: A Systematic Review and Meta-Analysis. Front Physiol [Internet]. 2018 Aug 7 [cited 2024 Jun 2];9. Available from: https://www.frontiersin.org/journals/physiology/articles/10.3389/fphys.2018.01057/full

68. Frank R, Larimore J. Yoga as a method of symptom management in multiple sclerosis. Front Neurosci [Internet]. 2015 Apr 30 [cited 2024 Jun 2];9. Available from: https://www.frontiersin.org/journals/neuroscience/articles/10.3389/fnins.2015.00133/full

69. Dhungana RR, Khatiwoda SR, Gurung Y, Pedišić Ž, de Courten M. Yoga for hypertensive patients: a study on barriers and facilitators of its implementation in primary care. Glob Health Action. 2021 Jan 1;14(1):1952753.

70. Nigg C, Weber C, Schipperijn J, Reichert M, Oriwol D, Worth A, et al. Urban-Rural Differences in Children’s and Adolescent’s Physical Activity and Screen-Time Trends Across 15 Years. Health Educ Behav. 2022 Oct 1;49(5):789–800.

71. Müller C, Paulsen L, Bucksch J, Wallmann-Sperlich B. Built and natural environment correlates of physical activity of adults living in rural areas: a systematic review. Int J Behav Nutr Phys Act. 2024 May 3;21(1):52.

72. Patel J, Katapally TR, Khadilkar A, Bhawra J. The interplay between air pollution, built environment, and physical activity: Perceptions of children and youth in rural and urban India. Health Place. 2024 Jan 1;85:103167.

73. Katapally TR, Rainham D, Muhajarine N. Factoring in weather variation to capture the influence of urban design and built environment on globally recommended levels of moderate to vigorous physical activity in children. BMJ Open. 2015 Nov 1;5(11):e009045.

74. Mitchell CA, Clark AF, Gilliland JA. Built Environment Influences of Children’s Physical Activity: Examining Differences by Neighbourhood Size and Sex. Int J Environ Res Public Health. 2016 Jan;13(1):130.

